# Early effectiveness of the BNT162b2 LP.8.1 vaccine against COVID-19 emergency department, urgent care, and outpatient visits in the US Veterans Affairs Healthcare System

**DOI:** 10.64898/2026.01.22.26344618

**Authors:** Haley J. Appaneal, Vrishali V. Lopes, Jennifer L. Nguyen, Hannah R. Volkman, Evan J. Zasowski, Aisling R. Caffrey

## Abstract

This study estimated early vaccine effectiveness (VE) of the BNT162b2 LP.8.1-adapted vaccine against emergency department/urgent care (ED/UC) and outpatient visits using a test-negative case-control design. Patients from the US Veterans Affairs Healthcare System with an acute respiratory infection (ARI) who underwent SARS-CoV-2 testing from September 10 – November 30, 2025 were included. VE was estimated using multivariable logistic regression adjusted for patient demographics and clinical characteristics. Among 34,455 ARI episodes, 10.7% were COVID-19 cases and 2.4% were vaccinated. BNT162b2 LP.8.1 vaccine was effective during the early 2025–2026 respiratory virus season. VE was 57% (95% confidence interval [CI] 39%–70%) against ED/UC visits and 54% (95% CI 15%–75%) against outpatient visits at approximately 4 weeks post-vaccination. These findings inform shared decision-making in clinical practice and support the continued importance of COVID-19 vaccination in populations for whom it is recommended.

## BACKGROUND

The 2025/2026 formulations of COVID-19 vaccines were adapted to target the SARS-CoV-2 sublineage LP.8.1 based on guidance from the US Food and Drug Administration (FDA) to match currently circulating strains.^1^ The US Centers for Disease Control and Prevention (CDC) recommends the 2025/2026 COVID-19 vaccine for individuals 6 months and older based on individual-based decision making, noting that vaccination is especially important for certain groups including those 65 years of age and older and those with high risk conditions as identified by the CDC.^2^ Real-world data from the past several years demonstrate the effectiveness of adapted mRNA COVID-19 vaccines in reducing the risk of mild-to-moderate (outpatient visits and emergency department/urgent care (ED/UC) and severe (hospitalization and mortality) COVID-19 outcomes.^3-10^ Despite guidance from public health agencies and infectious diseases experts on the continued importance of vaccination for avoiding severe COVID-19 illness,^2,11^ vaccination rates continue to decline. As of early December 2025, only 15.0% (95% confidence interval [CI] 13.9-16.2) of adults aged 18 years and older and 32.5% (95% CI 28.5-36.5) of adults 65 years and older received the 2025/2026 Formula COVID-19 vaccine.^11,12^ The availability of timely real-world effectiveness data for the LP.8.1–adapted vaccine plays an important role in informing clinical decision-making, reinforcing vaccination recommendations, and supporting vaccine uptake throughout the respiratory virus season.^13,14^ This study aimed to estimate the early vaccine effectiveness (VE) of the BNT162b2 LP.8.1-adapted vaccine against ED/UC and outpatient visits.

## METHODS

### Ethics Approval

This study complies with all applicable ethical regulations. The VA Providence Healthcare System (VAPHS) Institutional Review Board (IRB) determined that this study was exempt, and approval was granted by the VAPHS Research and Development Committee. Informed consent was not required for this retrospective study of existing health records.

### Design, Data Source

This study used a test-negative case-control design and analyzed clinical data from the national Veterans Affairs Healthcare System, which captures care delivered within the largest integrated healthcare system in the US. VA datasets integrate data from the VA’s electronic health record, including vaccine administration information (both within and outside the VA system), as well as detailed records of inpatient stays and outpatient visits.^15^

### Study Population and Setting

Adult patients (aged ≥18 years) diagnosed with an acute respiratory infection (ARI, **Supplemental Table 1**) and tested for SARS-CoV-2 by a nucleic acid amplification test (NAAT) or rapid antigen test (RAT) during a VA Healthcare System ED/UC or outpatient visit, without subsequent hospitalization, between September 10, 2025 and November 30, 2025 were included (**Supplemental Figure 1**). Similar to our previous work, corresponding SARS-CoV-2 testing had to occur between 14 days before and 3 days after the ARI encounter of interest.^5^ Several exclusion criteria were applied including: (1) no VA inpatient or outpatient visits in the prior year; (2) receipt of a 2025-2026 Formula COVID-19 vaccine other than the BNT162b2 LP.8.1 vaccine; (3) receipt of the BNT162b2 LP.8.1 vaccine with an unknown date of administration; (4) receipt of the BNT162b2 LP.8.1 vaccine within 14 days before the ARI encounter; (5) receipt of a 2024-2025 Formula COVID-19 vaccine (KP.2-or JN.1-adapted vaccine) <60 days before the ARI encounter; (6) receipt of SARS-CoV-2 antiviral treatment (nirmatrelvir/ritonavir, remdesivir, or molnupiravir) within 90 days before the ARI encounter; (7) receipt of SARS-CoV-2 monoclonal antibodies within 180 days before the ARI encounter; or (8) another ARI encounter in the previous 30 days. Patients could only contribute multiple ARI episodes if episodes were more than 30 days apart.

### Outcomes

Cases were defined as patients with a positive SARS-CoV-2 test and controls as those with a negative test result. The same definitions were used for each outcome category, ED/UC visits and outpatient visits.

### Exposure

Exposure was defined as receipt of the BNT162b2 LP.8.1 vaccine 14 or more days before the ARI episode. Those who did not receive a 2025-2026 Formula COVID-19 vaccine of any kind were considered unexposed.

### Statistical Analyses

VE was calculated by comparing the odds of BNT162b2 LP.8.1 vaccine receipt in SARS-CoV-2 positive cases and test-negative controls for each outcome category (ED/UC visits and outpatient visits). The following equation was used to calculate VE: (1-adjusted odds ratio) X 100. Odds ratios were estimated using multivariable logistic regression and adjusted for age category (18–64, 65–74, or ≥75 years), sex (male or female), race (Black, White, or other race), ethnicity (Hispanic or non-Hispanic), body mass index category (underweight, healthy weight, overweight, obese, or missing), Charlson Comorbidity Index (0, 1, 2, 3, or ≥4), receipt of pneumococcal vaccine in the past 5 years (yes or no), number of interactions with the VA healthcare system in the year prior (hospital admission, nursing home admission, ED/UC visit, primary care visit; 0 or ≥1 for each), prior documented SARS-CoV-2 infection (yes or no), smoking status (current/former smoker or never smoker/unknown), immunocompromised status (yes or no), and US Census region (Northeast, Midwest, South, or West), consistent with previous work.^4^ All logistic regression models were assessed for assumptions and model fit.

Receipt of the BNT162b2 LP.8.1 COVID-19 vaccine without the influenza vaccine was uncommon and precluded inclusion of influenza vaccination as a covariate in the adjusted model due to small and zero cell counts. As such, we performed a sensitivity analysis comparing those who received both the COVID-19 and influenza vaccines versus those who received neither. Those who received only the COVID-19 vaccine or only the influenza vaccine were excluded from this sensitivity analysis. We also conducted another sensitivity analysis excluding influenza positive controls. SAS (Version 9.4 and Enterprise Guide 8.3, SAS Institute Inc., Cary, NC, USA) was used for analyses

## DATA AVAILABILITY

Identifiable protected health data from the Veterans Health Administration support the findings of this study and cannot be made publicly available. Privacy regulations do not allow for open sharing of the individual-level data used in this study. After verifying de-identification, the Veterans Health Administration may approve sharing some of the study data but it may not include all final study data. Requests for such data should be made to the corresponding author and are subject to approvals by the ethics board, privacy office, and information systems and security office.

## RESULTS

From September 10 to November 30, 2025, 34,455 ARI episodes with a SARS-CoV-2 test in the VA ED/UC or outpatient setting were included (**Supplemental Figure 1**). Most (68.7%) episodes represented ED/UC visits and the remainder (31.3%) were outpatient visits. Patients aged 65 years and older accounted for 53.7% of the study population and most of those included were male (85.1%). More than one-third had a Charlson Comorbidity Index score of 3 or higher (36.2%) and immunocompromised status was common (41.3%). Overall, only 2.4% received the BNT162b2 LP.8.1 vaccine, with a median time since vaccination of 29 days (interquartile range 20-41). Among 3,696 cases and 30,759 controls, 1.2% and 2.6% received the BNT162b2 LP.8.1 vaccine, respectively.

Differences in demographic and clinical characteristics by case-control status are presented in **Table 1** and **Supplemental Table 2**. COVID-19 test-positive cases were more likely to have a Charlson Comorbidity Index score of 0 (37.6% *vs* 31.6%; *P*<.001) and less likely to have an immunocompromising condition (30.4% *vs* 42.6%; *P*<.001) compared to test-negative controls.. Demographic and clinical characteristics by case-control status for ED/UC visits are presented in **Supplemental Table 3** and for outpatient visits in **Supplemental Table 4**. Demographics and clinical characteristics by BNT162b2 LP.8.1 vaccination status are presented in **Supplemental Table 5**.

**Table 1.**
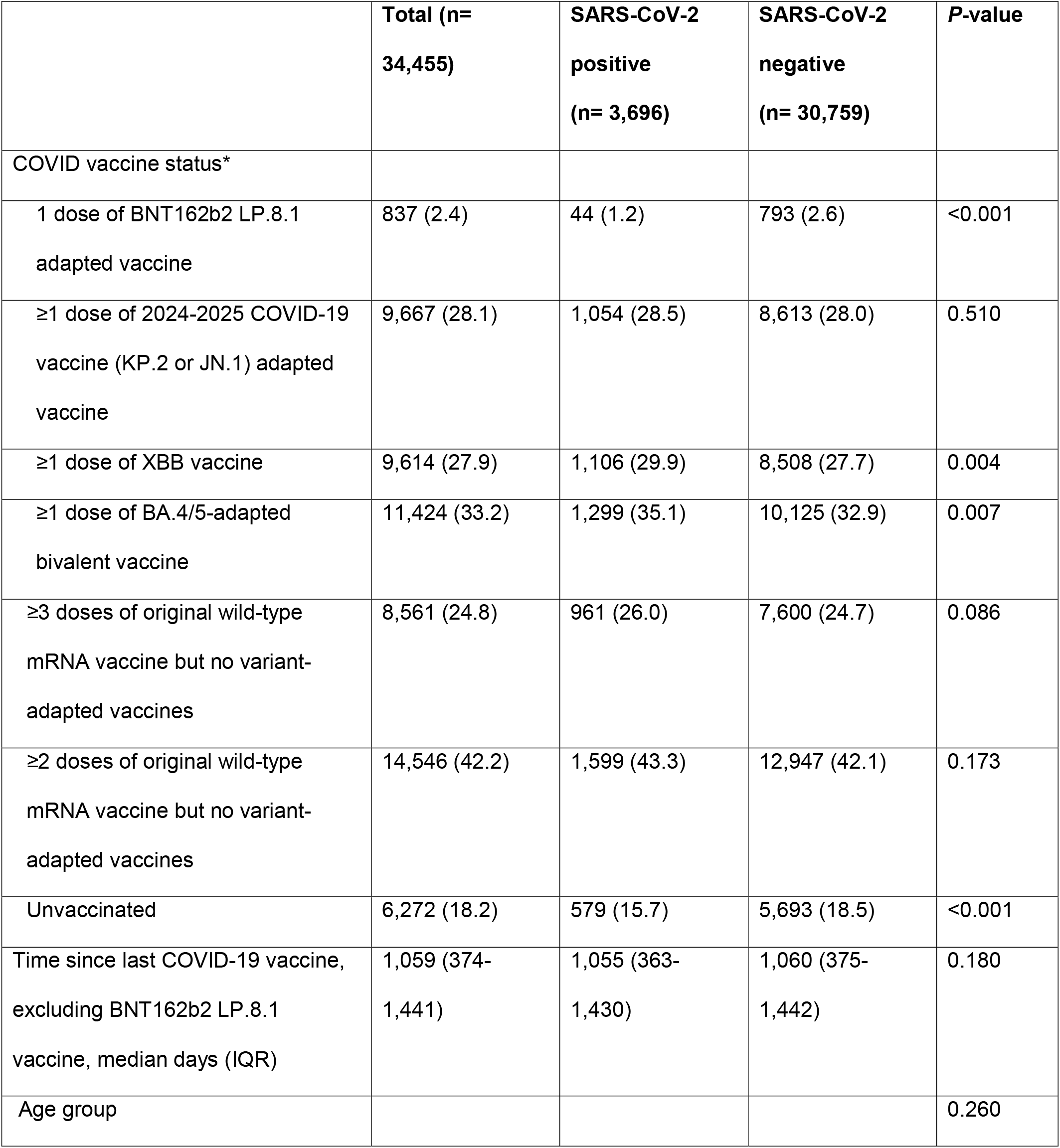
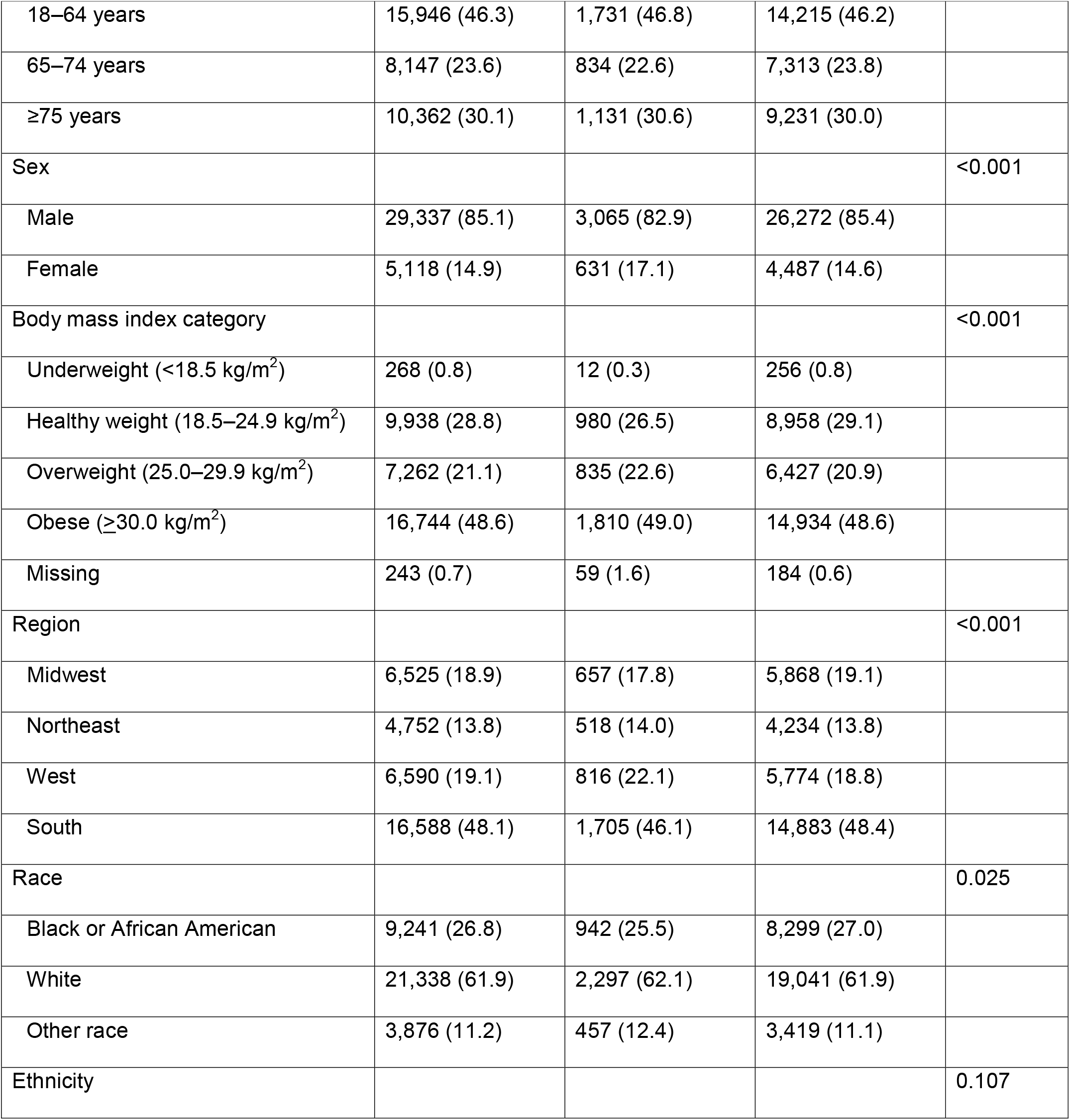
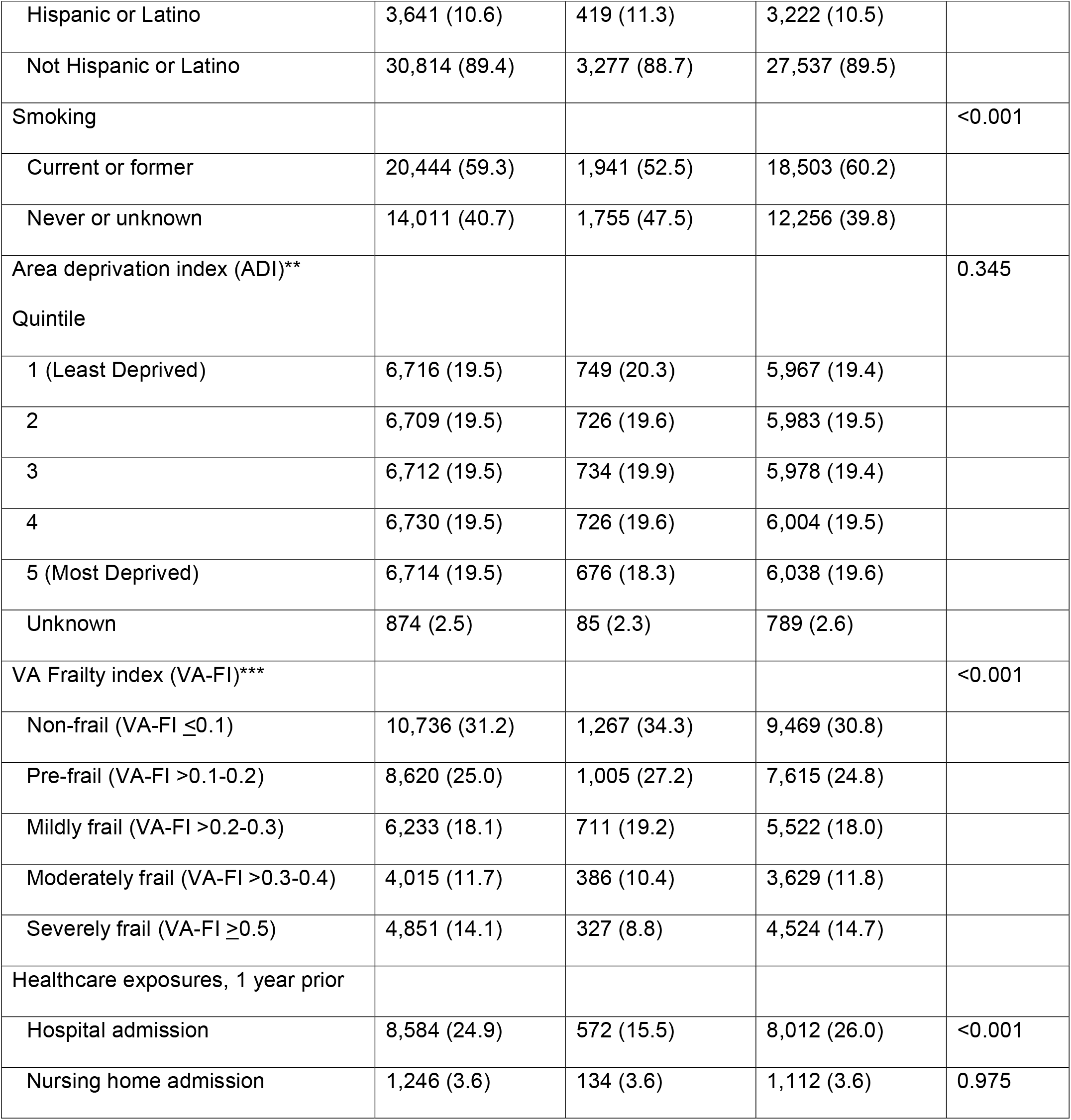
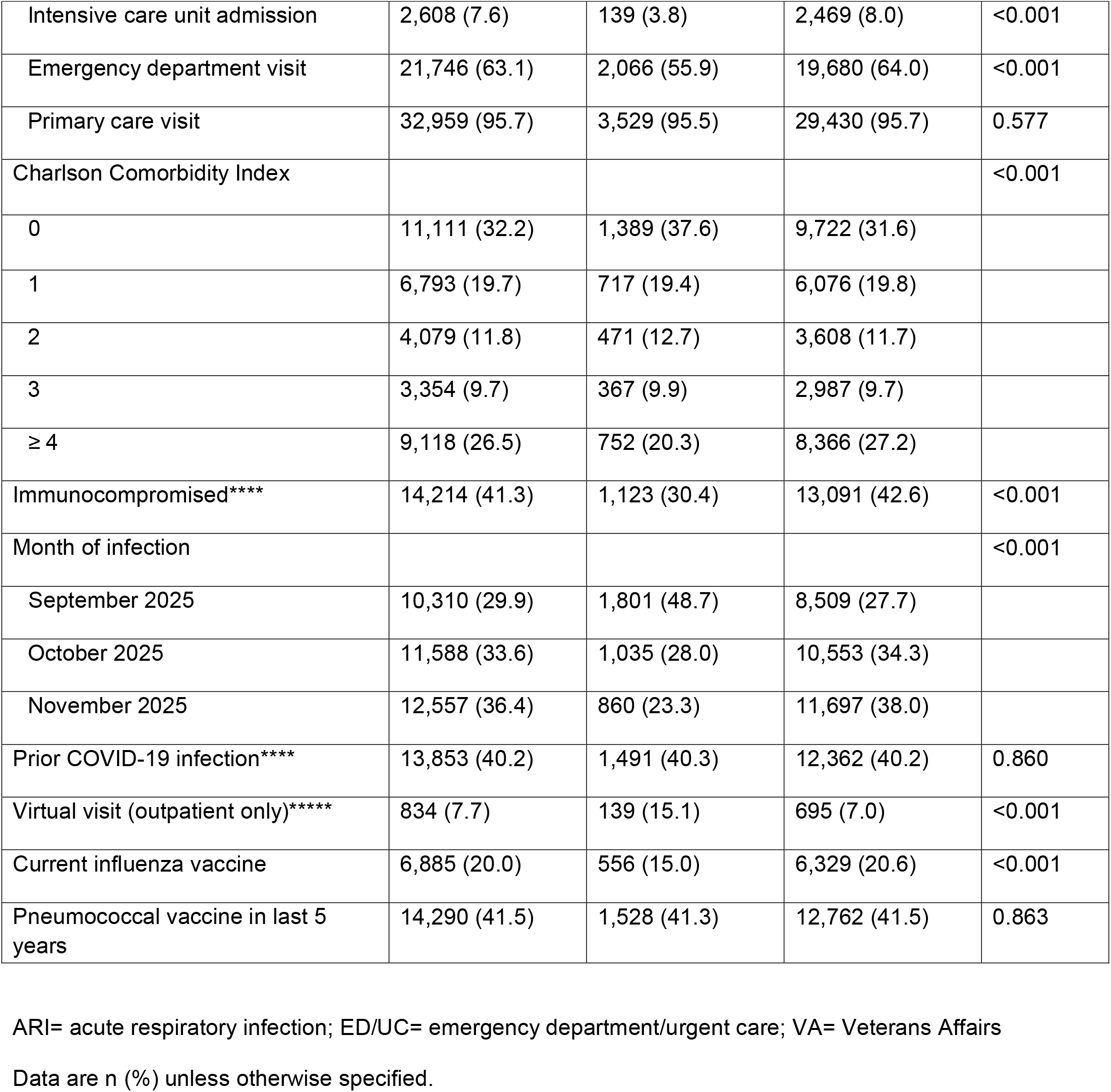
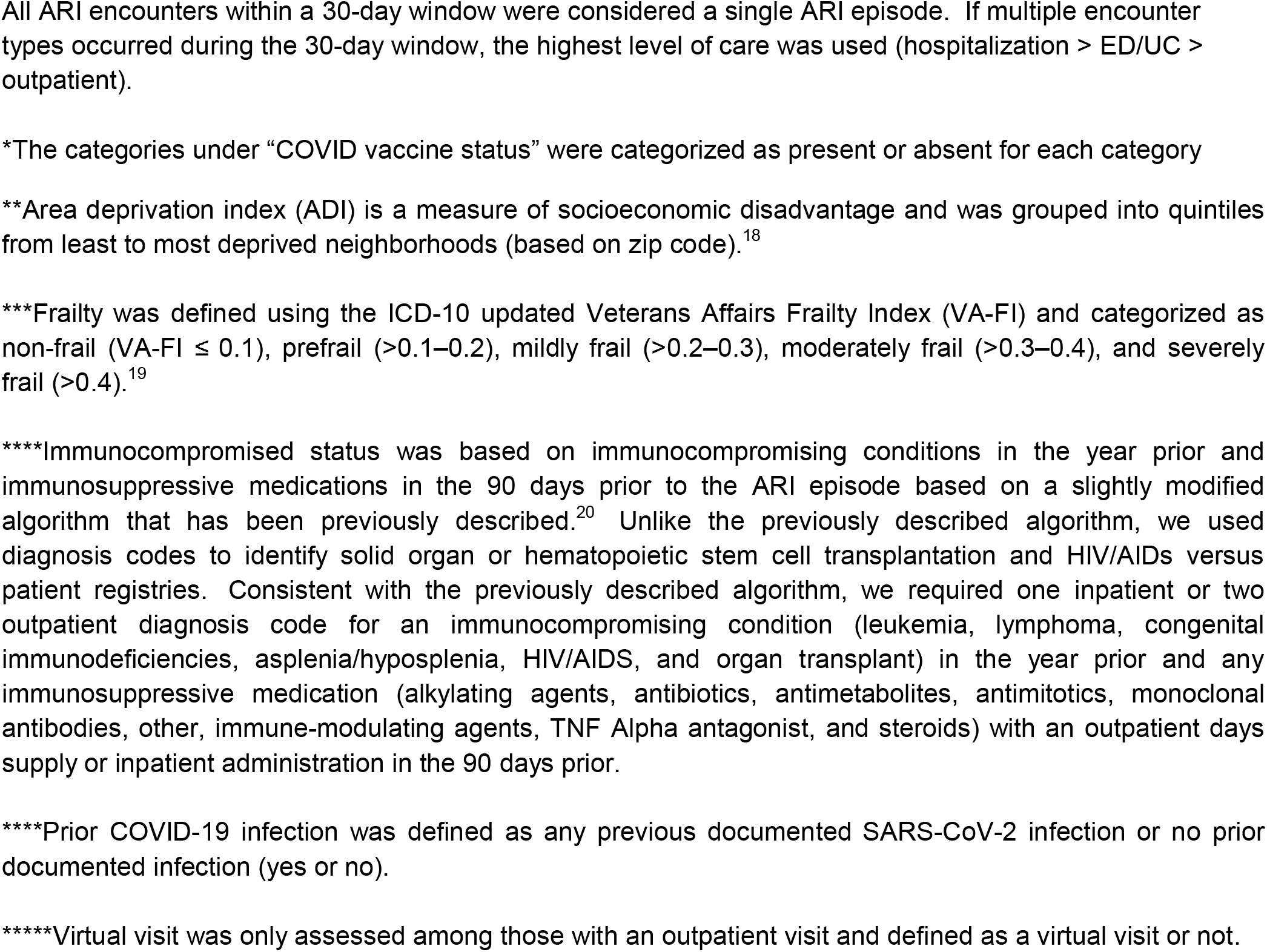
Demographics and clinical characteristics of acute respiratory infection episodes (ED/UC visits, outpatient visits) with SARS-CoV-2 testing by COVID-19 case-control status.

The overall adjusted VE of the BNT162b2 LP.8.1 vaccine, compared with no 2025–2026 Formula COVID-19 vaccination, was 56% (95% confidence interval [CI], 40%–68%; median [interquartile range (IQR), 29 [20–41] days since vaccination) against the composite outcome of ED/UC or outpatient visits. VE was 57% (95% CI, 39%–70%; median [IQR] 32 [21–42] days since dose) for ED/UC visits and 54% (95% CI, 15%–75%; median [IQR] 24 [16–37] days since dose) for outpatient visits. (**Figure 1**).

**Figure 1.**
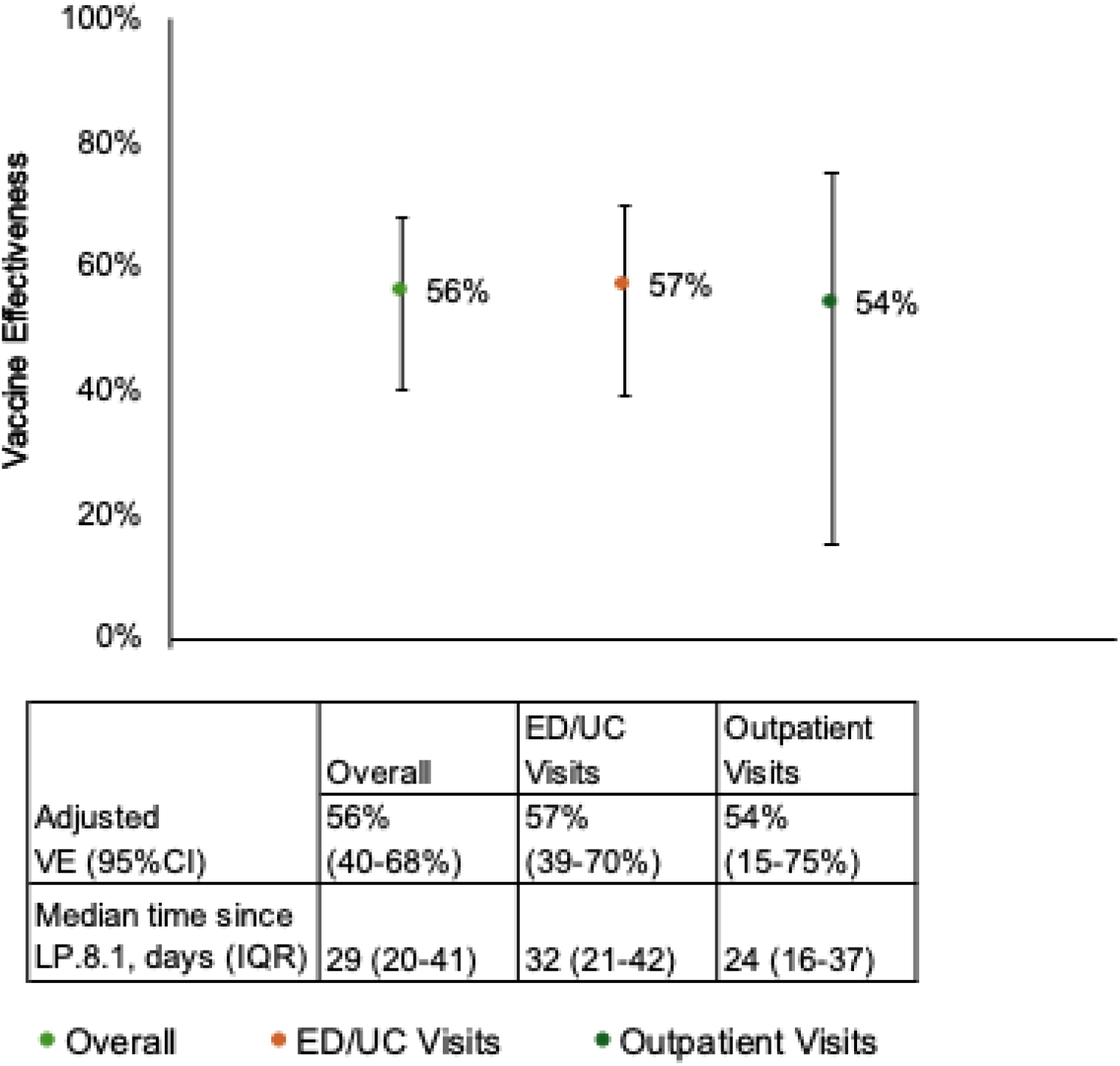
Adjusted effectiveness of the BNT162b2 LP.8.1 vaccine by COVID-19 outcomes ED/UC = emergency department/urgent care; IQR = interquartile range; LP.8.1 = BNT162b2 LP.8.1 adapted vaccine Compared the odds of receiving the 2025/2026 BNT162b2 LP.8.1 strain-adapted COVID-19 vaccine between SARS-CoV-2 positive cases and SARS-CoV-2 negative controls. Adjusted for age (18–64, 65–74, >75 years), sex (male or female), race (Black, White, or other race), ethnicity (Hispanic or non-Hispanic), body mass index (BMI) categories (underweight, healthy weight, overweight, obese, missing), Charlson Comorbidity Index (0, 1, 2, 3, ≥ 4), receipt of pneumococcal vaccine in the past 5 years (yes or no), hospital admission, nursing home admission, ED/UC visit, primary care visit; 0 or ≥1 for each), prior documented SARS-CoV-2 infection (yes or no), smoking status (current or former smoker or never smoker), immunocompromised (yes or no), and Census region (Northeast, Midwest, South, or West).

Results of sensitivity analyses among those who received both the COVID-19 and influenza vaccines were consistent with the primary analysis, with an adjusted VE of 64% (95% CI 47%–75%) against ED/UC visits and 59% (95% CI 24%–78%) against outpatient visits (**Supplemental Table 6**). Results of sensitivity analyses that excluded influenza positive controls were also consistent with the primary analysis with an adjusted VE of 56% (95% CI 38%–69%) against ED/UC visits and 53% (95% CI 14%–75%) against outpatient visits (**Supplemental Table 7**).

## DISCUSSION

This study presents early real-world effectiveness estimates for the BNT162b2 LP.8.1 vaccine against ED/UC and outpatient visits, and is one of the first real-world evaluations of the 2025–2026 Formula COVID-19 vaccine. The BNT162b2 LP.8.1 vaccine demonstrated 57% effectiveness against ED/UC visits and 54% effectiveness against outpatient visits at about one month since vaccination. These early estimates are comparable to early-season effectiveness observed for the BNT162b2 KP.2 vaccine during the 2024–2025 season (September 5–November 30, 2024), which showed effectiveness of 57% against ED/UC visits and 56% against outpatient visits.^4^

Only 2.4% of our VA cohort received the updated BNT162b2 LP.8.1 vaccine, which was lower than the previous year (3.7%).^4^ This pattern is consistent with CDC data demonstrating lower uptake of the 2025– 2026 Formula COVID-19 vaccine relative to the 2024–2025 Formula vaccine. By mid-December, uptake of the 2025–2026 Formula vaccine was lower than that of the 2024–2025 Formula vaccine among adults aged ≥18 years (15% vs 21%) and ≥65 years (32.5% vs 45%).^12,16^ Lower vaccination rates this year may be related to delays in updated CDC COVID-19 vaccine recommendations, as well as a shift from a universal recommendation for all individuals aged ≥6 months to guidance emphasizing individual-based clinical decision-making.^17^ In clinical discussions, it is important that patients and others involved in their care decisions understand that COVID-19 vaccines provide protection not only against severe disease but also against milder COVID-19 outcomes. Public health efforts should continue to emphasize the importance of COVID-19 vaccination, particularly among populations at increased risk for severe outcomes, including adults aged 65 years and older. Strategies such as risk-based outreach, clinician decision support tools, and expanded pharmacist-led vaccination programs may help improve acceptance and uptake of COVID-19 vaccines.

There are several limitations to this analysis, similar to our previous work.^4,5^ The test-negative case-control study design is susceptible to selection bias. VE may be underestimated because individuals with mild COVID-19 may be less likely to seek medical care and therefore would not be captured in the study. Although the regression models were adjusted for measured covariates, residual confounding due to unmeasured or unknown factors may persist. Similar to our analysis last year, we could not adjust for current season influenza vaccination because too few individuals received the LP.8.1 vaccine without the influenza vaccine resulting in unstable VE estimates. This study did not assess VE against hospitalization, as too few COVID-19 hospitalizations occurred among LP.8.1-vaccinated individuals during the analysis period. Given the demographic and clinical characteristics of the VA population, which is older, predominantly male, with a higher prevalence of chronic conditions, generalizability to the broader U.S. population may be limited.

In conclusion, VE of the BNT162b2 LP.8.1 vaccine was 57% against ED/UC visits and 54% against outpatient visits at about one month since vaccination, demonstrating real-world effectiveness during the early part of the 2025/2026 respiratory virus season against mild-to-moderate COVID-19 outcomes. Uptake of the BNT162b2 LP.8.1 vaccine remains low (2.4%), indicating a need for additional efforts to improve COVID-19 vaccination coverage during the remainder of the season, particularly among adults aged 65 years and older. Real-world VE should inform COVID-19 vaccination discussions during shared clinical decision-making between patients and healthcare providers.

## a. Funding

This study was conducted as a collaboration between the University of Rhode Island (URI), VA Providence Healthcare System, and Pfizer. Pfizer is the study sponsor. URI and VA Providence Healthcare System received funding from Pfizer in connection with the development of this manuscript and for data analysis.

## b. Other Acknowledgements

The views expressed are those of the authors and do not necessarily reflect the position or policy of the United States Department of Veterans Affairs.

The research study would not have been possible without the health information from patients under the care of the Veterans Health Administration. We express our gratitude to the VA patients for their invaluable contributions to medical and scientific progress.

## Author Contributions Statement

Conception and design of the study: Haley J. Appaneal, Vrishali V. Lopes, Laura Puzniak, Evan J. Zasowski, Aisling R. Caffrey

Data generation: Haley J. Appaneal, Vrishali V. Lopes, Aisling R. Caffrey

Analysis and/or interpretation of the data: Haley J. Appaneal, Vrishali V. Lopes, Laura Puzniak, Evan J. Zasowski, Aisling R. Caffrey

Preparation or critical revision of the manuscript: Haley J. Appaneal, Vrishali V. Lopes, Laura Puzniak, Evan J. Zasowski, Aisling R. Caffrey

## Competing Interests Statement

Haley J. Appaneal has received research funding from Pfizer and Merck.

Vrishali V. Lopes has no competing interest to declare.

Jennifer L. Nguyen, Hannah R. Volkman, and Evan J. Zasowski are employees and shareholders of Pfizer Inc.

Aisling R. Caffrey has received research funding from Pfizer.

