## Supplementary Material for "Early effectiveness of the BNT162b2 LP.8.1 vaccine against COVID-19 emergency department, urgent care, and outpatient visits in the US Veterans Affairs Healthcare System"

**Supplemental Figure 1.** Study selection criteria

Acute respiratory infection (ARI) encounters with a SARS-Cov-2 test in the VA from September 10, 2025 through November 30, 2025 without subsequest hospitalization (n=56,198)

Exclusions (N=21,743) No outpatient or inpatient visit in prior year (n=340) Received 2025-2026 COVID-19 vaccine other than BNT162b2 LP.8.1	vaccine (n=1,102) BNT162b2 LP.8.1 adapted vaccine administration date unknown (n=21) LP.8.1 vaccine received <14 days before the ARI encounter (n=844) Received 2024-2025 COVID-19 vaccine (KP.2 or JN.1) <60 days before	ARI encounter (n=138) SARS-CoV-2 antivirals received in the prior 90 days (n=3,006) SARS-CoV-2 monoclonal antibodies in the prior 180 days (n=0) ARI encounter ≤ 30 days from last day of previous episode (n=532) ARI encounter within 30 days considered single episode (n=15,760)

SARS-Cov-2 ARI episodes (n=34,455) ED/UC (n=23,662) Outpatient (n=10,793)

Test-positive COVID-19 cases n=3,696 (10.7%)

BNT162b2 xflp.8.1-adapted vaccine n=44 (1.2%)

No BNT162b2 LP.8.1-adapted vaccine	n=3,652 (98.8%)

Test-negative controls n=30,759 (89.3%)

BNT162b2 LP.8.1-adapted vaccine n=793 (2.6%)

No BNT162b2 LP.8.1-adapted vaccine	n=29,966 (97.4%)

ED/UC= emergency department/urgent care; VA = Veterans Affairs

Patients could contribute more than one ARI episode to the study if the episodes were more than 30 days apart.

**Supplemental Table 1.** Acute respiratory infection diagnosis codes (ICD-10)

| **ICD-10 Code** | **Diagnosis** |
| --- | --- |
| A22.1 | Pulmonary anthrax |
| A37.00 | Whooping cough due to *Bordetella pertussis* without pneumonia |
| A37.01 | Whooping cough due to *Bordetella pertussis* with pneumonia |
| A37.10 | Whooping cough due to *Bordetella parapertussis* without pneumonia |
| A37.11 | Whooping cough due to *Bordetella parapertussis* with pneumonia |
| A37.80 | Whooping cough due to other Bordetella species without pneumonia |
| A37.81 | Whooping cough due to other Bordetella species with pneumonia |
| A37.90 | Whooping cough, unspecified species without pneumonia |
| A37.91 | Whooping cough, unspecified species with pneumonia |
| A48.1 | Legionnaires' disease |
| B25.0 | Cytomegaloviral pneumonitis |
| B34.2 | Coronavirus infection, unspecified |
| B44.0 | Invasive pulmonary aspergillosis |
| B77.81 | Ascariasis pneumonia |
| B97.29 | Other coronavirus as the cause of diseases classified elsewhere |
| J00* | Acute nasopharyngitis [common cold] |
| J01* | Acute sinusitis |
| J02* | Acute pharyngitis |
| J03* | Acute tonsillitis |
| J04* | Acute laryngitis and tracheitis |
| J05* | Acute obstructive laryngitis [croup] and epiglottitis |
| J06* | Acute upper respiratory infections of multiple and unspecified sites |
| J09.X1 | Influenza due to novel influenza a virus with pneumonia |
| J09.X2 | Influenza due to identified novel influenza A virus with other respiratory manifestations |
| J09.X3 | Influenza due to identified novel influenza A virus with gastrointestinal manifestations |
| J09.X9 | Influenza due to identified novel influenza A virus with other manifestations |
| J10.00 | Influenza due to other identified influenza virus with pneumonia |
| J10.01 | Influenza due to other identified influenza virus with the same other identified influenza virus pneumonia |
| J10.08 | Influenza due to other identified influenza virus with other specified pneumonia |
| J10.1 | Influenza due to other identified influenza virus with other respiratory manifestations |
| J10.2 | Influenza due to other identified influenza virus with gastrointestinal manifestations |
| J10.8* | Influenza due to other identified influenza virus with other manifestations |
| J10.81 | Influenza due to other identified influenza virus with encephalopathy |
| J10.82 | Influenza due to other identified influenza virus with myocarditis |
| J10.83 | Influenza due to other identified influenza virus with otitis media |
| J10.89 | Influenza due to other identified influenza virus with other manifestations |
| J11.00 | Influenza due to unidentified influenza virus with pneumonia |
| J11.08 | Influenza due to unidentified influenza virus with specified pneumonia |
| J11.1 | Influenza due to unidentified influenza virus with other respiratory manifestations |
| J11.2 | Influenza due to unidentified influenza virus with gastrointestinal manifestations |
| J11.81 | Influenza due to unidentified influenza virus with encephalopathy |
| J11.82 | Influenza due to unidentified influenza virus with myocarditis |
| J11.83 | Influenza due to unidentified influenza virus with otitis media |
| J11.89 | Influenza due to unidentified influenza virus with other manifestations |
| J12.0 | Adenoviral pneumonia |
| J12.1 | Respiratory syncytial virus pneumonia |
| J12.2 | Parainfluenza virus pneumonia |
| J12.3 | Human metapneumovirus pneumonia |
| J12.81 | Pneumonia due to SARS-associated coronavirus |
| J12.82 | Pneumonia due to coronavirus disease 2019 |
| J12.89 | Other viral pneumonia |
| J12.9 | Viral pneumonia, unspecified |
| J13 | Pneumonia due to *Streptococcus pneumoniae* |
| J14 | Pneumonia due to *Hemophilus influenzae* |
| J15.0 | Pneumonia due to *Klebsiella pneumoniae* |
| J15.1 | Pneumonia due to Pseudomonas |
| J15.20 | Pneumonia due to Staphylococcus, unspecified |
| J15.211 | Pneumonia due to methicillin susceptible *Staphylococcus aureus* |
| J15.212 | Pneumonia due to methicillin resistant *Staphylococcus aureus* |
| J.15.29 | Pneumonia due to other Staphylococcus |
| J15.3 | Pneumonia due to Streptococcus, group b |
| J15.4 | Pneumonia due to other Streptococci |
| J15.5 | Pneumonia due to *Escherichia coli* |
| J15.6 | Pneumonia due to other aerobic gram-negative bacteria |
| J15.7 | Pneumonia due to *Mycoplasma pneumoniae* |
| J15.8 | Pneumonia due to other specified bacteria |
| J15.9 | Unspecified bacterial pneumonia |
| J16.0 | Chlamydial pneumonia |
| J16.8 | Pneumonia due to other specified infectious organisms |
| J17 | Pneumonia in diseases classified elsewhere |
| J18.0 | Bronchopneumonia, unspecified organism |
| J18.1 | Lobar pneumonia, unspecified organism |
| J18.2 | Hypostatic pneumonia, unspecified organism |
| J18.8 | Other pneumonia, unspecified organism |
| J18.9 | Pneumonia, unspecified organism |
| J20.0 | Acute bronchitis due to *Mycoplasma pneumoniae* |
| J20.1 | Acute bronchitis due to *Hemophilus influenzae* |
| J20.2 | Acute bronchitis due to Streptococcus |
| J20.3 | Acute bronchitis due to coxsackievirus |
| J20.4 | Acute bronchitis due to parainfluenza virus |
| J20.5 | Acute bronchitis due to respiratory syncytial virus |
| J20.6 | Acute bronchitis due to rhinovirus |
| J20.7 | Acute bronchitis due to echovirus |
| J20.8 | Acute bronchitis due to other specified organisms |
| J20.9 | Acute bronchitis, unspecified |
| J21.* | Acute bronchiolitis |
| J21.0 | Acute bronchiolitis due to respiratory syncytial virus |
| J21.1 | Acute bronchiolitis due to human metapneumovirus |
| J21.8 | Acute bronchiolitis due to other specified organisms |
| J21.9 | Acute bronchiolitis, unspecified |
| J22 | Unspecified acute lower respiratory infection |
| J80 | Acute respiratory distress syndrome |
| J96.00 | Acute respiratory failure unspecified whether with hypoxia or hypercapnia |
| J96.01 | Acute respiratory failure with hypoxia |
| J96.02 | Acute respiratory failure with hypercapnia |
| J96.10 | Chronic respiratory failure, unspecified with hypoxia or hypercapnia |
| J96.11 | Chronic respiratory failure with hypoxia |
| J96.12 | Chronic respiratory failure with hypercapnia |
| J96.20 | Acute and chr resp failure, unspecified with hypoxia or hypercapnia |
| J96.21 | Acute and chronic respiratory failure with hypoxia |
| J96.22 | Acute and chronic respiratory failure with hypercapnia |
| J96.90 | Respiratory failure, unspecified, unspecified with hypoxia or hypercapnia |
| J96.91 | Respiratory failure, unspecified with hypoxia |
| J96.92 | Respiratory failure, unspecified with hypercapnia |
| M35.81 | Multisystem inflammatory syndrome |
| R04.2 | Hemoptysis |
| R05 | Cough |
| R05.1 | Acute cough |
| R05.2 | Subacute cough |
| R05.3 | Chronic cough |
| R05.4 | Cough syncope |
| R05.8 | Other specified cough |
| R05.9 | Cough, unspecified |
| R06.00 | Dyspnea/abnormalities of breathing unspecified |
| R06.02 | Shortness of breath |
| R06.03 | Acute respiratory distress |
| R06.09 | Other forms of dyspnea |
| R06.1 | Stridor |
| R06.82 | Tachypnea, not elsewhere classified |
| R06.89 | Other abnormalities of breathing |
| R07.1 | Chest pain on breathing |
| R09.0* | Asphyxia and hypoxemia |
| R09.01 | Asphyxia |
| R09.02 | Hypoxemia |
| R09.1 | Pleurisy |
| R09.2 | Respiratory arrest |
| R50.9 | Fever, unspecified |
| U04* | SARS (WHO 2019) |
| u04.9 | SARS, unspecified (WHO 2019) |
| U07.1 | COVID-19 |
| U07.2 | COVID-19, virus not identified (clinically diagnosed) |

**Supplemental Table 2.** Medical history of acute respiratory infection episodes (ED/UC visits, outpatient visits) with SARS-CoV-2 testing, by COVID-19 case-control status

|  | **Total (n= 34,455)** | **SARS-CoV-2 positive**  **(n= 3,696)** | **SARS-CoV-2 negative**  **(n= 30,759)** | ***P*-value** |
| --- | --- | --- | --- | --- |
| Medical History |  |  |  |  |
| Acute cerebrovascular disease | 3,696 (10.7) | 317 (8.6) | 3,379 (11.0) | <0.001 |
| Acute myocardial infarction | 202 (0.6) | 16 (0.4) | 186 (0.6) | 0.196 |
| Alcohol and substance related disorders | 5,433 (15.8) | 462 (12.5) | 4,971 (16.2) | <0.001 |
| Any cancer or malignancy | 8,196 (23.8) | 829 (22.4) | 7,367 (24.0) | 0.040 |
| Aortic and peripheral arterial embolism or thrombosis | 45 (0.1) | <5 (<0.1) | 43 (0.1) | 0.173 |
| Asthma | 1,768 (5.1) | 161 (4.4) | 1,607 (5.2) | 0.024 |
| Benign prostatic hyperplasia | 4,892 (14.2) | 447 (12.1) | 4,445 (14.5) | <0.001 |
| Cardiac dysrhythmias | 5,111 (14.8) | 430 (11.6) | 4,681 (15.2) | <0.001 |
| Chronic kidney disease | 7,262 (21.1) | 692 (18.7) | 6,570 (21.4) | <0.001 |
| Chronic obstructive pulmonary disease and bronchiectasis | 3,353 (9.7) | 217 (5.9) | 3,136 (10.2) | <0.001 |
| Congestive heart failure | 5,347 (15.5) | 372 (10.1) | 4,975 (16.2) | <0.001 |
| Coronary artery disease | 3,037 (8.8) | 256 (6.9) | 2,781 (9.0) | <0.001 |
| Coronary atherosclerosis and other heart disease | 2,965 (8.6) | 247 (6.7) | 2,718 (8.8) | <0.001 |
| Delirium, dementia, and other cognitive disorders | 2,110 (6.1) | 142 (3.8) | 1,968 (6.4) | <0.001 |
| Diabetes with or without chronic complications | 11,949 (34.7) | 1,189 (32.2) | 10,760 (35.0) | <0.001 |
| Epilepsy | 289 (0.8) | 19 (0.5) | 270 (0.9) | 0.022 |
| Human immunodeficiency virus (HIV) infection | 259 (0.8) | 20 (0.5) | 239 (0.8) | 0.117 |
| Hypertension | 21,743 (63.1) | 2,192 (59.3) | 19,551 (63.6) | <0.001 |
| Influenza | 345 (1.0) | 27 (0.7) | 318 (1.0) | 0.080 |
| Liver diseases | 3,526 (10.2) | 329 (8.9) | 3,197 (10.4) | 0.005 |
| Mental health conditions | 17,819 (51.7) | 1,837 (49.7) | 15,982 (52.0) | 0.009 |
| Osteoarthritis | 4,772 (13.8) | 504 (13.6) | 4,268 (13.9) | 0.691 |
| Peripheral and visceral atherosclerosis | 2,608 (7.6) | 224 (6.1) | 2,384 (7.8) | <0.001 |
| Pneumonia | 2,637 (7.7) | 140 (3.8) | 2,497 (8.1) | <0.001 |
| Pulmonary heart disease | 1,448 (4.2) | 92 (2.5) | 1,356 (4.4) | <0.001 |
| Rheumatoid arthritis | 553 (1.6) | 74 (2.0) | 479 (1.6) | 0.042 |
| Septicemia | 559 (1.6) | 32 (0.9) | 527 (1.7) | <0.001 |
| Thyroid disorder | 1,042 (3.0) | 116 (3.1) | 926 (3.0) | 0.668 |
| Tuberculosis | 17 (0.0) | <5 (<0.1) | 17 (0.1) | 0.153 |

ARI= acute respiratory infection; ED/UC= emergency department/urgent care; VA= Veterans Affairs

Data are n (%) unless otherwise specified.

All ARI encounters within a 30-day window were considered a single ARI episode. If multiple encounter types occurred during the 30-day window, the highest level of care was used (ED/UC > outpatient).

Medical history included underlying conditions and diagnoses in the year prior to the ARI episode, identified using international classification of diseases (ICD)-10 codes.

**Supplemental Table 3.** Demographics and clinical characteristics of acute respiratory infection ED/UC visits with SARS-CoV-2 testing, by COVID-19 case-control status

|  | **Total (n=23,662)** | **SARS-CoV-2 positive**  **(n=2,773)** | **SARS-CoV-2 negative**  **(n=20,889)** | ***P*-value** |
| --- | --- | --- | --- | --- |
| COVID vaccine status* |  |  |  |  |
| 1 dose of BNT162b2 LP.8.1 adapted vaccine | 563 (2.4) | 33 (1.2) | 530 (2.5) | <0.001 |
| ≥1 dose of 2024-2025 COVID-19 vaccine (KP.2 or JN.1) adapted vaccine | 6,384 (27.0) | 788 (28.4) | 5,596 (26.8) | 0.070 |
| ≥1 dose of XBB vaccine | 6,368 (26.9) | 838 (30.2) | 5,530 (26.5) | <0.001 |
| ≥1 dose of BA.4/5-adapted bivalent vaccine | 7,647 (32.3) | 986 (35.6) | 6,661 (31.9) | <0.001 |
| ≥3 doses of original wild-type mRNA vaccine but no variant-adapted vaccines | 5,900 (24.9) | 735 (26.5) | 5,165 (24.7) | 0.042 |
| ≥2 doses of original wild-type mRNA vaccine but no variant-adapted vaccines | 10,128 (42.8) | 1,231 (44.4) | 8,897 (42.6) | 0.072 |
| Unvaccinated | 4,371 (18.5) | 404 (14.6) | 3,967 (19.0) | <0.001 |
| Time since last COVID-19 vaccine, excluding BNT162b2 LP.8.1 vaccine, median days (IQR) | 1,079 (380-1,447) | 1,062 (365-1,433) | 1,082 (382-1,449) | 0.021 |
| Age group |  |  |  | 0.006 |
| 18–64 years | 11,579 (48.9) | 1,295 (46.7) | 10,284 (49.2) |  |
| 65–74 years | 5,464 (23.1) | 633 (22.8) | 4,831 (23.1) |  |
| ≥75 years | 6,619 (28.0) | 845 (30.5) | 5,774 (27.6) |  |
| Sex |  |  |  | 0.008 |
| Male | 20,081 (84.9) | 2,306 (83.2) | 17,775 (85.1) |  |
| Female | 3,581 (15.1) | 467 (16.8) | 3,114 (14.9) |  |
| Body mass index category |  |  |  | 0.004 |
| Underweight (<18.5 kg/m^2^) | 138 (0.6) | 7 (0.3) | 131 (0.6) |  |
| Healthy weight (18.5–24.9 kg/m^2^) | 6,397 (27.0) | 726 (26.2) | 5,671 (27.1) |  |
| Overweight (25.0–29.9 kg/m^2^) | 5,094 (21.5) | 641 (23.1) | 4,453 (21.3) |  |
| Obese (≥30.0 kg/m^2^) | 11,856 (50.1) | 1,369 (49.4) | 10,487 (50.2) |  |
| Missing | 177 (0.7) | 30 (1.1) | 147 (0.7) |  |
| Region |  |  |  | <0.001 |
| Midwest | 4,409 (18.6) | 462 (16.7) | 3,947 (18.9) |  |
| Northeast | 3,346 (14.1) | 404 (14.6) | 2,942 (14.1) |  |
| West | 4,758 (20.1) | 656 (23.7) | 4,102 (19.6) |  |
| South | 11,149 (47.1) | 1,251 (45.1) | 9,898 (47.4) |  |
| Race |  |  |  | 0.338 |
| Black or African American | 6,644 (28.1) | 746 (26.9) | 5,898 (28.2) |  |
| White | 14,356 (60.7) | 1,708 (61.6) | 12,648 (60.5) |  |
| Other race | 2,662 (11.3) | 319 (11.5) | 2,343 (11.2) |  |
| Ethnicity |  |  |  | 0.090 |
| Hispanic or Latino | 2,535 (10.7) | 323 (11.6) | 2,212 (10.6) |  |
| Not Hispanic or Latino | 21,127 (89.3) | 2,450 (88.4) | 18,677 (89.4) |  |
| Smoking |  |  |  | <0.001 |
| Current or former | 13,572 (57.4) | 1,430 (51.6) | 12,142 (58.1) |  |
| Never or unknown | 10,090 (42.6) | 1,343 (48.4) | 8,747 (41.9) |  |
| Area deprivation index (ADI)** Quintile |  |  |  | 0.237 |
| 1 (Least Deprived) | 4,717 (19.9) | 572 (20.6) | 4,145 (19.8) |  |
| 2 | 4,687 (19.8) | 546 (19.7) | 4,141 (19.8) |  |
| 3 | 4,611 (19.5) | 562 (20.3) | 4,049 (19.4) |  |
| 4 | 4,564 (19.3) | 544 (19.6) | 4,020 (19.2) |  |
| 5 (Most Deprived) | 4,392 (18.6) | 481 (17.3) | 3,911 (18.7) |  |
| Unknown | 691 (2.9) | 68 (2.5) | 623 (3.0) |  |
| VA Frailty index (VA-FI)*** |  |  |  | <0.001 |
| Non-frail (VA-FI *≤*0.1) | 8,088 (34.2) | 964 (34.8) | 7,124 (34.1) |  |
| Pre-frail (VA-FI >0.1-0.2) | 6,143 (26.0) | 753 (27.2) | 5,390 (25.8) |  |
| Mildly frail (VA-FI >0.2-0.3) | 4,273 (18.1) | 535 (19.3) | 3,738 (17.9) |  |
| Moderately frail (VA-FI >0.3-0.4) | 2,524 (10.7) | 289 (10.4) | 2,235 (10.7) |  |
| Severely frail (VA-FI ≥0.5) | 2,634 (11.1) | 232 (8.4) | 2,402 (11.5) |  |
| Healthcare exposures, 1 year prior |  |  |  |  |
| Hospital admission | 4,491 (19.0) | 415 (15.0) | 4,076 (19.5) | <0.001 |
| Nursing home admission | 525 (2.2) | 39 (1.4) | 486 (2.3) | 0.002 |
| Intensive care unit admission | 1,041 (4.4) | 83 (3.0) | 958 (4.6) | <0.001 |
| Emergency department visit | 15,536 (65.7) | 1,708 (61.6) | 13,828 (66.2) | <0.001 |
| Primary care visit | 22,708 (96.0) | 2,674 (96.4) | 20,034 (95.9) | 0.188 |
| Charlson Comorbidity Index |  |  |  | 0.001 |
| 0 | 8,334 (35.2) | 1,047 (37.8) | 7,287 (34.9) |  |
| 1 | 4,872 (20.6) | 538 (19.4) | 4,334 (20.7) |  |
| 2 | 2,794 (11.8) | 350 (12.6) | 2,444 (11.7) |  |
| 3 | 2,280 (9.6) | 280 (10.1) | 2,000 (9.6) |  |
| ≥ 4 | 5,382 (22.7) | 558 (20.1) | 4,824 (23.1) |  |
| Immunocompromised**** | 10,035 (42.4) | 889 (32.1) | 9,146 (43.8) | <0.001 |
| Month of infection |  |  |  | <0.001 |
| September 2025 | 6,587 (27.8) | 1,292 (46.6) | 5,295 (25.3) |  |
| October 2025 | 7,979 (33.7) | 802 (28.9) | 7,177 (34.4) |  |
| November 2025 | 9,096 (38.4) | 679 (24.5) | 8,417 (40.3) |  |
| Prior COVID-19 infection**** | 9,837 (41.6) | 1,154 (41.6) | 8,683 (41.6) | 0.961 |
| Current influenza vaccine | 4,711 (19.9) | 444 (16.0) | 4,267 (20.4) | <0.001 |
| Pneumococcal vaccine in last 5 years | 9,639 (40.7) | 1,147 (41.4) | 8,492 (40.7) | 0.475 |

ARI= acute respiratory infection; ED/UC= emergency department/urgent care; VA= Veterans Affairs

Data are n (%) unless otherwise specified.

All ARI encounters within a 30-day window were considered a single ARI episode. If multiple encounter types occurred during the 30-day window, the highest level of care was used (hospitalization > ED/UC > outpatient).

*The categories under “COVID vaccine status” were categorized as present or absent for each category

**Area deprivation index (ADI) is a measure of socioeconomic disadvantage and was grouped into quintiles from least to most deprived neighborhoods (based on zip code).^20^

***Frailty was defined using the ICD-10 updated Veterans Affairs Frailty Index (VA-FI) and categorized as non-frail (VA-FI ≤ 0.1), prefrail (>0.1–0.2), mildly frail (>0.2–0.3), moderately frail (>0.3–0.4), and severely frail (>0.4).^21^

****Immunocompromised status was based on immunocompromising conditions in the year prior and immunosuppressive medications in the 90 days prior to the ARI episode based on a slightly modified algorithm that has been previously described.^12^  Unlike the previously described algorithm, we used diagnosis codes to identify solid organ or hematopoietic stem cell transplantation and HIV/AIDs versus patient registries.  Consistent with the previously described algorithm, we required one inpatient or two outpatient diagnosis code for an immunocompromising condition (leukemia, lymphoma, congenital immunodeficiencies, asplenia/hyposplenia, HIV/AIDS, and organ transplant) in the year prior and any immunosuppressive medication (alkylating agents, antibiotics, antimetabolites, antimitotics, monoclonal antibodies, other, immune-modulating agents, TNF Alpha antagonist, and steroids) with an outpatient days supply or inpatient administration in the 90 days prior.

****Prior COVID-19 infection was defined as any previous documented SARS-CoV-2 infection or no prior documented infection (yes or no).

**Supplemental Table 4.** Demographics and clinical characteristics of acute respiratory infection outpatient visits with SARS-CoV-2 testing, by COVID-19 case-control status

|  | **Total (n=10,793)** | **SARS-CoV-2 positive**  **(n=923)** | **SARS-CoV-2 negative**  **(n=9,870)** | ***P*-value** |
| --- | --- | --- | --- | --- |
| COVID vaccine status* |  |  |  |  |
| 1 dose of BNT162b2 LP.8.1 adapted vaccine | 274 (2.5) | 11 (1.2) | 263 (2.7) | 0.007 |
| ≥1 dose of 2024-2025 COVID-19 vaccine (KP.2 or JN.1) adapted vaccine | 3,283 (30.4) | 266 (28.8) | 3,017 (30.6) | 0.270 |
| ≥1 dose of XBB vaccine | 3,246 (30.1) | 268 (29.0) | 2,978 (30.2) | 0.472 |
| ≥1 dose of BA.4/5-adapted bivalent vaccine | 3,777 (35.0) | 313 (33.9) | 3,464 (35.1) | 0.470 |
| ≥3 doses of original wild-type mRNA vaccine but no variant-adapted vaccines | 2,661 (24.7) | 226 (24.5) | 2,435 (24.7) | 0.901 |
| ≥2 doses of original wild-type mRNA vaccine but no variant-adapted vaccines | 4,418 (40.9) | 368 (39.9) | 4,050 (41.0) | 0.492 |
| Unvaccinated | 1,901 (17.6) | 175 (19.0) | 1,726 (17.5) | 0.261 |
| Time since last COVID-19 vaccine, excluding BNT162b2 LP.8.1 vaccine, median days (IQR) | 987 (364-1,425) | 1,029 (357-1,427.5) | 979 (364-1,425) | 0.703 |
| Age group |  |  |  | <0.001 |
| 18–64 years | 4,367 (40.5) | 436 (47.2) | 3,931 (39.8) |  |
| 65–74 years | 2,683 (24.9) | 201 (21.8) | 2,482 (25.1) |  |
| ≥75 years | 3,743 (34.7) | 286 (31.0) | 3,457 (35.0) |  |
| Sex |  |  |  | 0.001 |
| Male | 9,256 (85.8) | 759 (82.2) | 8,497 (86.1) |  |
| Female | 1,537 (14.2) | 164 (17.8) | 1,373 (13.9) |  |
| Body mass index category |  |  |  | <0.001 |
| Underweight (<18.5 kg/m^2^) | 130 (1.2) | 5 (0.5) | 125 (1.3) |  |
| Healthy weight (18.5–24.9 kg/m^2^) | 3,541 (32.8) | 254 (27.5) | 3,287 (33.3) |  |
| Overweight.0 (25–29.9 kg/m^2^) | 2,168 (20.1) | 194 (21.0) | 1,974 (20.0) |  |
| Obese (≥30.0 kg/m^2^) | 4,888 (45.3) | 441 (47.8) | 4,447 (45.1) |  |
| Missing | 66 (0.6) | 29 (3.1) | 37 (0.4) |  |
| Region |  |  |  | 0.590 |
| Midwest | 2,116 (19.6) | 195 (21.1) | 1,921 (19.5) |  |
| Northeast | 1,406 (13.0) | 114 (12.4) | 1,292 (13.1) |  |
| West | 1,832 (17.0) | 160 (17.3) | 1,672 (16.9) |  |
| South | 5,439 (50.4) | 454 (49.2) | 4,985 (50.5) |  |
| Race |  |  |  | 0.001 |
| Black or African American | 2,597 (24.1) | 196 (21.2) | 2,401 (24.3) |  |
| White | 6,982 (64.7) | 589 (63.8) | 6,393 (64.8) |  |
| Other race | 1,214 (11.2) | 138 (15.0) | 1,076 (10.9) |  |
| Ethnicity |  |  |  | 0.872 |
| Hispanic or Latino | 1,106 (10.2) | 96 (10.4) | 1,010 (10.2) |  |
| Not Hispanic or Latino | 9,687 (89.8) | 827 (89.6) | 8,860 (89.8) |  |
| Smoking |  |  |  | <0.001 |
| Current or former | 6,872 (63.7) | 511 (55.4) | 6,361 (64.4) |  |
| Never or unknown | 3,921 (36.3) | 412 (44.6) | 3,509 (35.6) |  |
| Area deprivation index (ADI)** Quintile |  |  |  | 0.948 |
| 1 (Least Deprived) | 1,999 (18.5) | 177 (19.2) | 1,822 (18.5) |  |
| 2 | 2,022 (18.7) | 180 (19.5) | 1,842 (18.7) |  |
| 3 | 2,101 (19.5) | 172 (18.6) | 1,929 (19.5) |  |
| 4 | 2,166 (20.1) | 182 (19.7) | 1,984 (20.1) |  |
| 5 (Most Deprived) | 2,322 (21.5) | 195 (21.1) | 2,127 (21.6) |  |
| Unknown | 183 (1.7) | 17 (1.8) | 166 (1.7) |  |
| VA Frailty index (VA-FI)*** |  |  |  | <0.001 |
| Non-frail (VA-FI *≤*0.1) | 2,648 (24.5) | 303 (32.8) | 2,345 (23.8) |  |
| Pre-frail (VA-FI >0.1-0.2) | 2,477 (23.0) | 252 (27.3) | 2,225 (22.5) |  |
| Mildly frail (VA-FI >0.2-0.3) | 1,960 (18.2) | 176 (19.1) | 1,784 (18.1) |  |
| Moderately frail (VA-FI >0.3-0.4) | 1,491 (13.8) | 97 (10.5) | 1,394 (14.1) |  |
| Severely frail (VA-FI ≥0.5) | 2,217 (20.5) | 95 (10.3) | 2,122 (21.5) |  |
| Healthcare exposures, 1 year prior |  |  |  |  |
| Hospital admission | 4,093 (37.9) | 157 (17.0) | 3,936 (39.9) | <0.001 |
| Nursing home admission | 721 (6.7) | 95 (10.3) | 626 (6.3) | <0.001 |
| Intensive care unit admission | 1,567 (14.5) | 56 (6.1) | 1,511 (15.3) | <0.001 |
| Emergency department visit | 6,210 (57.5) | 358 (38.8) | 5,852 (59.3) | <0.001 |
| Primary care visit | 10,251 (95.0) | 855 (92.6) | 9,396 (95.2) | <0.001 |
| Charlson Comorbidity Index |  |  |  | <0.001 |
| 0 | 2,777 (25.7) | 342 (37.1) | 2,435 (24.7) |  |
| 1 | 1,921 (17.8) | 179 (19.4) | 1,742 (17.6) |  |
| 2 | 1,285 (11.9) | 121 (13.1) | 1,164 (11.8) |  |
| 3 | 1,074 (10.0) | 87 (9.4) | 987 (10.0) |  |
| ≥ 4 | 3,736 (34.6) | 194 (21.0) | 3,542 (35.9) |  |
| Immunocompromised**** | 4,179 (38.7) | 234 (25.4) | 3,945 (40.0) | <0.001 |
| Month of infection |  |  |  | <0.001 |
| September 2025 | 3,723 (34.5) | 509 (55.1) | 3,214 (32.6) |  |
| October 2025 | 3,609 (33.4) | 233 (25.2) | 3,376 (34.2) |  |
| November 2025 | 3,461 (32.1) | 181 (19.6) | 3,280 (33.2) |  |
| Prior COVID-19 infection**** | 4,016 (37.2) | 337 (36.5) | 3,679 (37.3) | 0.646 |
| Virtual visit (outpatient only)***** | 834 (7.7) | 139 (15.1) | 695 (7.0) | <0.001 |
| Current influenza vaccine | 2,174 (20.1) | 112 (12.1) | 2,062 (20.9) | <0.001 |
| Pneumococcal vaccine in last 5 years | 4,651 (43.1) | 381 (41.3) | 4,270 (43.3) | 0.244 |

ARI= acute respiratory infection; ED/UC= emergency department/urgent care; VA= Veterans Affairs

Data are n (%) unless otherwise specified.

All ARI encounters within a 30-day window were considered a single ARI episode. If multiple encounter types occurred during the 30-day window, the highest level of care was used (hospitalization > ED/UC > outpatient).

*The categories under “COVID vaccine status” were categorized as present or absent for each category

**Area deprivation index (ADI) is a measure of socioeconomic disadvantage and was grouped into quintiles from least to most deprived neighborhoods (based on zip code).^20^

***Frailty was defined using the ICD-10 updated Veterans Affairs Frailty Index (VA-FI) and categorized as non-frail (VA-FI ≤ 0.1), prefrail (>0.1–0.2), mildly frail (>0.2–0.3), moderately frail (>0.3–0.4), and severely frail (>0.4).^21^

****Immunocompromised status was based on immunocompromising conditions in the year prior and immunosuppressive medications in the 90 days prior to the ARI episode based on a slightly modified algorithm that has been previously described.^12^  Unlike the previously described algorithm, we used diagnosis codes to identify solid organ or hematopoietic stem cell transplantation and HIV/AIDs versus patient registries.  Consistent with the previously described algorithm, we required one inpatient or two outpatient diagnosis code for an immunocompromising condition (leukemia, lymphoma, congenital immunodeficiencies, asplenia/hyposplenia, HIV/AIDS, and organ transplant) in the year prior and any immunosuppressive medication (alkylating agents, antibiotics, antimetabolites, antimitotics, monoclonal antibodies, other, immune-modulating agents, TNF Alpha antagonist, and steroids) with an outpatient days supply or inpatient administration in the 90 days prior.

****Prior COVID-19 infection was defined as any previous documented SARS-CoV-2 infection or no prior documented infection (yes or no).

*****Virtual visit was only assessed among those with an outpatient visit and defined as a virtual visit or not.

**Supplemental Table 5.** Demographics and clinical characteristics of acute respiratory infection episodes (ED/UC visits, outpatient visits) with SARS-CoV-2 testing, by vaccination status

| **Variable** | **Total (n=34,455)** | **Received BNT162b2 LP.8.1 vaccine (n=837)** | **No LP.8.1 vaccine of any kind (n=33,618)** | ***P*-value** |
| --- | --- | --- | --- | --- |
| COVID vaccine status |  |  |  |  |
| ≥1 dose of 2024-2025 COVID-19 vaccine (KP.2 or JN.1) adapted vaccine | 9,667 (28.1) | 707 (84.5) | 8,960 (26.7) | <0.001 |
| ≥1 dose of XBB vaccine | 9,614 (27.9) | 665 (79.5) | 8,949 (26.6) | <0.001 |
| ≥1 dose of BA.4/5-adapted bivalent vaccine | 11,424 (33.2) | 677 (80.9) | 10,747 (32.0) | <0.001 |
| ≥3 doses of original wild-type mRNA vaccine but no variant-adapted vaccines | 8,561 (24.8) | 122 (14.6) | 8,439 (25.1) | <0.001 |
| ≥2 doses of original wild-type mRNA vaccine but no variant-adapted vaccines | 14,546 (42.2) | 140 (16.7) | 14,406 (42.9) | <0.001 |
| Unvaccinated | 6,272 (18.2) | <5 (<0.6) | 6,269 (18.6) | <0.001 |
| Time since last COVID-19 vaccine, excluding BNT162b2 LP.8.1 vaccine, median days (IQR) | 1,059 (374-1,441) | 397 (362-422) | 1,078 (375-1,446) | <0.001 |
| Age group |  |  |  | <0.001 |
| 18–64 years | 15,946 (46.3) | 135 (16.1) | 15,811 (47.0) |  |
| 65–74 years | 8,147 (23.6) | 246 (29.4) | 7,901 (23.5) |  |
| ≥75 years | 10,362 (30.1) | 456 (54.5) | 9,906 (29.5) |  |
| Sex |  |  |  | <0.001 |
| Male | 29,337 (85.1) | 778 (93.0) | 28,559 (85.0) |  |
| Female | 5,118 (14.9) | 59 (7.0) | 5,059 (15.0) |  |
| Body mass index category |  |  |  | 0.027 |
| Underweight (<18.5 kg/m^2^) | 268 (0.8) | 5 (0.6) | 263 (0.8) |  |
| Healthy weight (18.5–24.9 kg/m^2^) | 9,938 (28.8) | 269 (32.1) | 9,669 (28.8) |  |
| Overweight (25.0–29.9 kg/m^2^) | 7,262 (21.1) | 179 (21.4) | 7,083 (21.1) |  |
| Obese (≥30.0 kg/m^2^) | 16,744 (48.6) | 384 (45.9) | 16,360 (48.7) |  |
| Missing | 243 (0.7) | <5 (<0.6) | 243 (0.7) |  |
| Region |  |  |  | <0.001 |
| Midwest | 6,525 (18.9) | 185 (22.1) | 6,340 (18.9) |  |
| Northeast | 4,752 (13.8) | 134 (16.0) | 4,618 (13.7) |  |
| West | 6,590 (19.1) | 196 (23.4) | 6,394 (19.0) |  |
| South | 16,588 (48.1) | 322 (38.5) | 16,266 (48.4) |  |
| Race |  |  |  | 0.564 |
| Black or African American | 9,241 (26.8) | 223 (26.6) | 9,018 (26.8) |  |
| White | 21,338 (61.9) | 529 (63.2) | 20,809 (61.9) |  |
| Other race | 3,876 (11.2) | 85 (10.2) | 3,791 (11.3) |  |
| Ethnicity |  |  |  | <0.001 |
| Hispanic or Latino | 3,641 (10.6) | 50 (6.0) | 3,591 (10.7) |  |
| Not Hispanic or Latino | 30,814 (89.4) | 787 (94.0) | 30,027 (89.3) |  |
| Smoking |  |  |  | 0.002 |
| Current or former | 20,444 (59.3) | 540 (64.5) | 19,904 (59.2) |  |
| Never or unknown | 14,011 (40.7) | 297 (35.5) | 13,714 (40.8) |  |
| Area deprivation index (ADI) Quintile |  |  |  | <0.001 |
| 1 (Least Deprived) | 6,716 (19.5) | 212 (25.3) | 6,504 (19.3) |  |
| 2 | 6,709 (19.5) | 175 (20.9) | 6,534 (19.4) |  |
| 3 | 6,712 (19.5) | 167 (20.0) | 6,545 (19.5) |  |
| 4 | 6,730 (19.5) | 140 (16.7) | 6,590 (19.6) |  |
| 5 (Most Deprived) | 6,714 (19.5) | 133 (15.9) | 6,581 (19.6) |  |
| Missing | 874 (2.5) | 10 (1.2) | 864 (2.6) |  |
| VA Frailty index (VA-FI)* |  |  |  | <0.001 |
| Non-frail (VA-FI *≤*0.1) | 10,736 (31.2) | 132 (15.8) | 10,604 (31.5) |  |
| Pre-frail (VA-FI >0.1-0.2) | 8,620 (25.0) | 194 (23.2) | 8,426 (25.1) |  |
| Mildly frail (VA-FI >0.2-0.3) | 6,233 (18.1) | 180 (21.5) | 6,053 (18.0) |  |
| Moderately frail (VA-FI >0.3-0.4) | 4,015 (11.7) | 136 (16.2) | 3,879 (11.5) |  |
| Severely frail (VA-FI ≥0.5) | 4,851 (14.1) | 195 (23.3) | 4,656 (13.8) |  |
| Healthcare exposures, 1 year prior |  |  |  |  |
| Hospital admission | 8,584 (24.9) | 281 (33.6) | 8,303 (24.7) | <0.001 |
| Nursing home admission | 1,246 (3.6) | 37 (4.4) | 1,209 (3.6) | 0.207 |
| Intensive care unit admission | 2,608 (7.6) | 59 (7.0) | 2,549 (7.6) | 0.565 |
| Emergency department visit | 21,746 (63.1) | 554 (66.2) | 21,192 (63.0) | 0.062 |
| Primary care visit | 32,959 (95.7) | 825 (98.6) | 32,134 (95.6) | <0.001 |
| Charlson Comorbidity Index |  |  |  |  |
| 0 | 11,111 (32.2) | 132 (15.8) | 10,979 (32.7) | <0.001 |
| 1 | 6,793 (19.7) | 149 (17.8) | 6,644 (19.8) |  |
| 2 | 4,079 (11.8) | 124 (14.8) | 3,955 (11.8) |  |
| 3 | 3,354 (9.7) | 96 (11.5) | 3,258 (9.7) |  |
| ≥ 4 | 9,118 (26.5) | 336 (40.1) | 8,782 (26.1) |  |
| Immunocompromised** | 14,214 (41.3) | 385 (46.0) | 13,829 (41.1) | 0.005 |
| Medical History*** |  |  |  |  |
| Acute cerebrovascular disease | 3,696 (10.7) | 143 (17.1) | 3,553 (10.6) | <0.001 |
| Acute myocardial infarction | 202 (0.6) | 14 (1.7) | 188 (0.6) | <0.001 |
| Alcohol and substance related disorders | 5,433 (15.8) | 122 (14.6) | 5,311 (15.8) | 0.338 |
| Any cancer or malignancy | 8,196 (23.8) | 272 (32.5) | 7,924 (23.6) | <0.001 |
| Aortic and peripheral arterial embolism or thrombosis | 45 (0.1) | <5 (<0.6) | 43 (0.1) | 0.380 |
| Asthma | 1,768 (5.1) | 55 (6.6) | 1,713 (5.1) | 0.056 |
| Benign prostatic hyperplasia | 4,892 (14.2) | 182 (21.7) | 4,710 (14.0) | <0.001 |
| Cardiac dysrhythmias | 5,111 (14.8) | 206 (24.6) | 4,905 (14.6) | <0.001 |
| Chronic kidney disease | 7,262 (21.1) | 242 (28.9) | 7,020 (20.9) | <0.001 |
| Chronic obstructive pulmonary disease and bronchiectasis | 3,353 (9.7) | 112 (13.4) | 3,241 (9.6) | <0.001 |
| Congestive heart failure | 5,347 (15.5) | 188 (22.5) | 5,159 (15.3) | <0.001 |
| Coronary artery disease | 3,037 (8.8) | 107 (12.8) | 2,930 (8.7) | <0.001 |
| Coronary atherosclerosis and other heart disease | 2,965 (8.6) | 104 (12.4) | 2,861 (8.5) | <0.001 |
| Delirium, dementia, and other cognitive disorders | 2,110 (6.1) | 74 (8.8) | 2,036 (6.1) | <0.001 |
| Diabetes with or without chronic complications | 11,949 (34.7) | 355 (42.4) | 11,594 (34.5) | <0.001 |
| Epilepsy | 289 (0.8) | 9 (1.1) | 280 (0.8) | 0.448 |
| Human immunodeficiency virus (HIV) infection | 259 (0.8) | 9 (1.1) | 250 (0.7) | 0.273 |
| Hypertension | 21,743 (63.1) | 660 (78.9) | 21,083 (62.7) | <0.001 |
| Influenza | 345 (1.0) | 7 (0.8) | 338 (1.0) | 0.627 |
| Liver diseases | 3,526 (10.2) | 81 (9.7) | 3,445 (10.2) | 0.591 |
| Mental health conditions | 17,819 (51.7) | 354 (42.3) | 17,465 (52.0) | <0.001 |
| Osteoarthritis | 4,772 (13.8) | 161 (19.2) | 4,611 (13.7) | <0.001 |
| Peripheral and visceral atherosclerosis | 2,608 (7.6) | 109 (13.0) | 2,499 (7.4) | <0.001 |
| Pneumonia | 2,637 (7.7) | 95 (11.4) | 2,542 (7.6) | <0.001 |
| Pulmonary heart disease | 1,448 (4.2) | 51 (6.1) | 1,397 (4.2) | 0.006 |
| Rheumatoid arthritis | 553 (1.6) | 22 (2.6) | 531 (1.6) | 0.017 |
| Septicemia | 559 (1.6) | 12 (1.4) | 547 (1.6) | 0.662 |
| Thyroid disorder | 1,042 (3.0) | 32 (3.8) | 1,010 (3.0) | 0.172 |
| Tuberculosis | 17 (0.0) | <5 (<0.6) | 16 (0.0) | 0.355 |
| Month of infection |  |  |  | <0.001 |
| September 2025 | 10,310 (29.9) | 15 (1.8) | 10,295 (30.6) |  |
| October 2025 | 11,588 (33.6) | 178 (21.3) | 11,410 (33.9) |  |
| November 2025 | 12,557 (36.4) | 644 (76.9) | 11,913 (35.4) |  |
| Prior COVID-19 infection | 13,853 (40.2) | 340 (40.6) | 13,513 (40.2) | 0.804 |
| Virtual visit (outpatient only) | 834 (7.7) | 27 (9.9) | 807 (7.7) | 0.182 |
| Current influenza vaccine | 6,885 (20.0) | 781 (93.3) | 6,104 (18.2) | <0.001 |
| Pneumococcal vaccine in last 5 years | 14,290 (41.5) | 547 (65.4) | 13,743 (40.9) | <0.001 |

ARI= acute respiratory infection; ED/UC= emergency department/urgent care; VA= Veterans Affairs

Data are n (%) unless otherwise specified. Chi-square or Fisher’s Exact tests were used to compare differences in proportions between the groups. For continuous variables, comparisons were performed using the Wilcoxon Rank Sum test or a Student’s t-test, depending on the distribution of the data for the given variable.

All ARI encounters within a 30-day window were considered a single ARI episode. If multiple encounter types occurred during the 30-day window, the highest level of care was used (hospitalization > ED/UC > outpatient).

*VA Frailty index was categorized as non-frail (VA-FI ≤ 0.1), prefrail (>0.1–0.2), mildly frail (>0.2–0.3), moderately frail (>0.3–0.4), and severely frail (>0.4)

**Immunocompromised status was based on immunocompromising conditions in the year prior and immunosuppressive medications in the 90 days prior to the ARI episode based on a slightly modified algorithm that has been previously described. (Tartof SY, et al. Lancet Reg Health Am. 2022:9:100198.) Unlike the previously described algorithm, we used diagnosis codes to identify solid organ or hematopoietic stem cell transplantation and HIV/AIDs versus patient registries.  Consistent with the previously described algorithm, we required one inpatient or two outpatient diagnosis code for an immunocompromising condition (leukemia, lymphoma, congenital immunodeficiencies, asplenia/hyposplenia, HIV/AIDS, and organ transplant) in the year prior and any immunosuppressive medication (alkylating agents, antibiotics, antimetabolites, antimitotics, monoclonal antibodies, other, immune-modulating agents, TNF Alpha antagonist, and steroids) with an outpatient days supply or inpatient administration in the 90 days prior.

***Medical history included underlying conditions and diagnoses in the year prior to the ARI episode, identified using international classification of diseases (ICD)-10 codes.

**Supplemental Table 6.** Adjusted vaccine effectiveness of the BNT162b2 LP.8.1 vaccine and influenza vaccine compared with neither vaccination for ED/UC visits and outpatient visits

| **Outcome** | **Adjusted VE for COVID-19 and influenza vaccination compared with neither (N = 28,295)*** | |
| --- | --- | --- |
|  | **VE (95% CI)** | **Median time since LP.8.1 vaccine, days (IQR)** |
| Overall | 62 (47-72) | 29 (20-41) |
| ED/UC visits | 64 (47-75) | 31 (21-42) |
| Outpatient visits | 59 (24-78) | 24 (17-38) |

ED/UC = emergency department/urgent care; IQR = interquartile range; LP.8.1 = BNT162b2 LP.8.1 adapted vaccine; VE= vaccine effectiveness

*Analysis excludes those vaccinated against COVID but not influenza (n=56), and those vaccinated for influenza but not COVID (n=6,104).

Compared the odds of receiving the 2025/2026 BNT162b2 LP.8.1 strain-adapted COVID-19 vaccine and the influenza vaccine compared to neither vaccination between SARS-CoV-2 positive cases and SARS-CoV-2 negative controls. Adjusted for age (18–64, 65–74, >75 years), sex (male or female), race (Black, White, or other race), ethnicity (Hispanic or non-Hispanic), body mass index (BMI) categories (underweight, healthy weight, overweight, obese, missing), Charlson Comorbidity Index (0, 1, 2, 3, ≥ 4), receipt of pneumococcal vaccine in the past 5 years (yes or no), hospital admission, nursing home admission, ED/UC visit, primary care visit; 0 or ≥1 for each), prior documented SARS-CoV-2 infection (yes or no), smoking status (current/former smoker or never smoker/unknown), immunocompromised (yes or no), and Census region (Northeast, Midwest, South, or West).

**Supplemental Table 7.** Adjusted vaccine effectiveness of the BNT162b2 LP.8.1 vaccine for ED/UC visits and outpatient visits excluding influenza positive controls

| **Outcome** | **Adjusted VE for COVID-19 excluding influenza positive controls (N = 33,657)*** | |
| --- | --- | --- |
|  | **VE (95% CI)** | **Median time since LP.8.1 vaccine, days (IQR)** |
| Overall | 55 (39-67) | 29 (20-41) |
| ED/UC visits | 56 (38-69) | 32 (22-42) |
| Outpatient visits | 53 (14-75) | 24 (16-38) |

ED/UC = emergency department/urgent care; IQR = interquartile range; LP.8.1 = BNT162b2 LP.8.1 adapted vaccine; VE= vaccine effectiveness

*Analysis excludes influenza positive controls (n=798)

Compared the odds of receiving the 2025/2026 BNT162b2 LP.8.1 strain-adapted COVID-19 vaccine between SARS-CoV-2 positive cases and SARS-CoV-2 negative controls excluding influenza positive controls. Adjusted for age (18–64, 65–74, >75 years), sex (male or female), race (Black, White, or other race), ethnicity (Hispanic or non-Hispanic), body mass index (BMI) categories (underweight, healthy weight, overweight, obese, missing), Charlson Comorbidity Index (0, 1, 2, 3, ≥ 4), receipt of pneumococcal vaccine in the past 5 years (yes or no), hospital admission, nursing home admission, ED/UC visit, primary care visit; 0 or ≥1 for each), prior documented SARS-CoV-2 infection (yes or no), smoking status (current/former smoker or never smoker/unknown), immunocompromised (yes or no), and Census region (Northeast, Midwest, South, or West).
